# PyTMLE: A Flexible Python Library for Targeted Estimation of Survival and Competing Risks using Causal Machine Learning

**DOI:** 10.1101/2025.07.02.25330730

**Authors:** Jannis Guski, Mohamed Aborageh, Holger Fröhlich

**Affiliations:** Department of Bioinformatics, Fraunhofer Institute for Algorithms and Scientific Computing (SCAI), Schloss Birlinghoven, Sankt Augustin, 53757, Germany; Bonn-Aachen International Center for Information Technology (b-it), University of Bonn, Friedrich-Hirzebruch-Allee 6, Bonn, 53115, Germany

**Keywords:** Targeted Maximum Likelihood Estimation (TMLE), Competing Risks, Survival Analysis, Machine Learning

## Abstract

**Background:** Targeted estimation offers a robust and unbiased approach for causal inference of the average treatment effect (ATE) from observational data, even with confounding, dependent censoring, and competing risks. Its advantages include double robustness, statistical rigor, and flexible data-adaptive modeling, potentially leveraging machine/deep learning. However, existing implementations lack model selection flexibility and are R-based, hindering adoption by the Python-focused machine learning community.

**Results:** We propose `PyTMLE`, a flexible Python package for causal machine learning-based targeted estimation with survival outcomes and competing risks. `PyTMLE` supports `scikit-survival` and `pycox`, and inbuilt robustness checks based on *E-values*. `PyTMLE` is easy to use with initial estimation of nuisance parameters that are obtained via super learning by default. We showcase its basic usage on the established Hodgkin’s disease dataset, where our package reveals the protective effect of chemotherapy on relapse risk.

**Conclusions:** This package promotes targeted estimation in time-to-event analysis for applied machine learning, enabling fully data-adaptive nuisance parameter estimation, potentially with deep learning. Future enhancements may include time-dependent confounders and dynamic treatment regimes.

## 1 Background

Many biomedical and epidemiological studies aim to estimate how treatments or exposures influence the risk of events, like disease onset, over time. Estimating time-to-event outcomes is often complicated by right censoring, where the event of interest occurs outside the study period for some participants, and competing risks, where other events preclude the primary event. While right censoring has been addressed in survival analysis for decades, competing risks require specialized methods. Consequently, many Machine Learning (ML) techniques have been adapted for survival and competing risks analysis, including penalized Cox regression [1], random survival forests [2, 3], gradient boosting [4–6], and, more recently, deep learning models like DeepHit [7], DeepSurv [8], and Nnet-survival [9]. These methods enable flexible and data-adaptive analyses in time-to-event settings.

Estimating treatment effects can be biased by confounding and dependent censoring. For example, in observational studies, treatment assignment may be influenced by confounders, such as age and comorbidities, if a drug is preferentially prescribed to older, sicker patients. Similarly, dependent censoring can occur when patient characteristics, including treatment-related side effects, influence study dropout. In this case, censoring is not random and can bias estimates of the Average Treatment Effects (ATEs).

Targeted Maximum Likelihood Estimation (TMLE) addresses biases by integrating ML with semiparametric efficiency theory to yield robust and asymptotically efficient estimators for parameters of interest [10]. In causal inference with binary treatments, these parameters commonly represent counterfactual means under universal treatment or no treatment, from which the ATE can be derived. TMLE involves two stages: initial estimation of nuisance parameters followed by a targeting step that solves the Efficient Influence Curves (EICs) equation to refine these estimates [10]. When initial estimators converge rapidly (e.g., through cross-fitting or super learning [11]), the TMLE estimator becomes asymptotically linear, allowing for inference based on asymptotic normality [12].

A specific version of TMLE for survival and competing risks analysis with continuously distributed event times was proposed by Rytgaard & van der Laan [12]. In this particular TMLE framework, the parameters of interest in the update step are counterfactual conditional average cause-specific hazards. Additionally, propensity scores and the conditional censoring survival function are estimated as nuisance parameters. Like previous time-to-event versions of TMLE, estimation is doubly robust, meaning that the TMLE point estimate is consistent if either a) propensities and censoring survival are consistently estimated or b) conditional hazards are consistently estimated for confounding and censoring [13]. Chen et al. [14] implemented the method in the R library `concrete`.

TMLE provides robust, theoretically well founded causal inference through flexible, machine learning-based methods suitable for observational data and complex, highdimensional settings. While statistically rigorous TMLE methods are often developed in R, Python is the dominant language in applied ML and Artificial Intelligence (AI), evidenced by its top rankings in the TIOBE index [15] and GitHub Octoverse [16], and its prevalence among data scientists [17]. Although interoperability between Python and R exists via packages like `rpy2` [18] and `reticulate` [19], these solutions can be cumbersome, especially for complex models and deep learning applications.

We present `PyTMLE`, a Python package for TMLE of survival and competing risks estimands with binary treatments and baseline confounders. It is based on Rytgaard & van der Laan’s [12] methodology and its implementation in `concrete` [14]. `PyTMLE` provides maximum flexibility in the choice of model for the estimation of nuisance parameters in the first TMLE stage, thereby making it more adaptive to any given data than previous implementations. By default, all initial estimates are derived from a super learner design, which is a continuous stacking classifier for propensity scores, and a version of the discrete *state learner* proposed by Munch & Gerds [20] for conditional event and censoring hazards. The package should further encourage the use of targeted estimation in the applied ML and AI community.

### 1.1 Related software packages

Our package uses a translation of the R package `concrete` [14] into Python. While `concrete` isn’t the first R library for TMLE in survival analysis [21–23], no equivalent Python implementation exists. Existing TMLE options in Python packages like `zEpid` [24] and Uber’s CausalML [25] are limited to classification or regression and do not handle right censoring.

Python offers several ML libraries for survival analysis, notably pycox [26], `scikit-survival` [27], and `lifelines` [28]. `pycox` provides torch implementations of deep learning models, including *DeepSurv* [8] and the competing risks-aware *DeepHit* [7]. `scikit-survival`and lifelines focus on traditional models like Cox regression [29], random survival forests [2], and gradient boosting [4, 5]. However, these models generally lack support for competing risks with covariates, although `hazardous` [6] offers a gradient boosting extension for competing risks.

### 1.2 Benefits and relevance of the new package

Three key assets of `PyTMLE` can be emphasized to underline the relevance of the package for future biomedical and epidemiological research:

1. It unlocks the full potential of TMLE by pairing the statistical rigor of the targeted update step with fully flexible estimation of nuisance parameters. Unlike previous software, `PyTMLE` enables the inclusion of arbitrary cross-fitted ML models in a discrete super learner to get initial estimates, including pytorch-based deep learning in particular.
2. It bridges the gap between the statistics and (applied) ML / AI communities by bringing TMLE for survival and competing risks estimands to Python. Thereby, a broader group of data scientists and epidemiologists should be encouraged to perform causal inference on real-world data in the presence of right censoring.
3. It provides tools to assess to what extent basic assumptions for causal inference may be violated. In the absence of a ground truth for causal estimands, a close monitoring of assumptions and sensitivity analysis is crucial to determine the validity of predicted effects.

### 1.3 Outline of our manuscript

In Section 2, we describe the implementation of `PyTMLE`, including the underlying methodologies and the software architecture that enables flexible targeted estimation for survival and competing risks analysis. Section 3 demonstrates the application of `PyTMLE`through a case study using real-world biomedical data, highlighting its performance and practical utility. Finally, Section 4 concludes the manuscript by summarizing the benefits of PyTMLE, and outlining potential directions for future development.

## 2 Implementation

In the following, the TMLE framework by Rytgaard & van der Laan [12] will be recapitulated and our implementation presented. While remaining close to Chen et al.’s [14] version in general, there are some differences:

- The first TMLE stage is more flexible, allowing arbitrary ML models to be plugged into a discrete super learner pipeline. In principal, initial estimates can even be computed completely outside of the package and then passed directly to the second TMLE stage.
- Instead of only cause-specific Cox-type models, our super learner utilizes a library with Cox regression, random survival forests and DeepHit by default. This is possible because the super learner is based on the model-agnostic state learner [20], and facilitates an out-of-the-box application of the package.
- For the sake of simplicity, only deterministic interventions are supported (not intent-to-treat analysis).
- Confidence Intervals (CIs) cannot only be based on the closed-form variance computation derived from the EIC, but also on a non-parametric bootstrap of the second TMLE stage.
- Sensitivity analysis is available in the form of *E-values* [30] to assess how robust effect estimates are with regard to unobserved confounding.

### 2.1 Causal Inference Framework

We will first define our causal estimands and then transform them to statistical estimands that can be estimated from observed data.

The causal estimands will be derived from counterfactual risks of having encountered an event of interest by a given time. Our implementation provides for counterfactuals from binary interventions *a^*^ ∈* {0, 1} within the framework of a design with two treatment arms, i.e., treated vs. untreated. The counterfactual competing risks data for intervention *a^*^* can then be described as a tuple

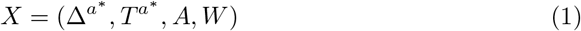

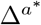 and 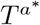 denote the counterfactual index of the event (out of all *J* competing events) which would be encountered first under intervention *a^*^*, and the counterfactual time until that event would occur, respectively. If this was event *j*, then 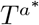 would equal the event time 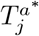, and if no event was observed in the follow-up period, 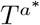 would equal the censoring time 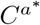. *A ∈* {0, 1} is the observed treatment, and *W* are the observed baseline covariates.

The counterfactual risk of having experienced event *j* at target time *t_k_* under intervention *d* can be expressed as probabilities

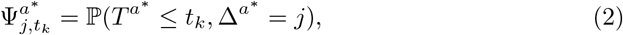

From these we can compose our causal estimands, which are ATEs defined in terms of either Risk Differences (RDs) or Risk Ratios (RRs):

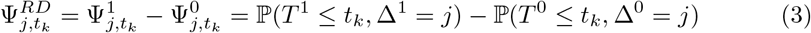

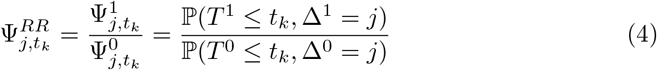

If we further define the conditional censoring survival probability at time *t* as *G*(*t a, w*) = ℙ(*C > t | A* = *a, W* = *w*) and propensity scores as *π*(*a | w*) = ℙ(*A* = *a | W* = *w*), the causal estimands can be identified from observed data if the following assumptions are met [12]:

- *Positivity* There is a non-zero probability of remaining under observation during follow-up for each intervention level: *∀a^*^* : *G*(*t_k_ | a^*^, W*)*π*(*a^*^ | W*) *>* 0.
- *Conditional exchangeability* Given the observed treatment and covariates, the observed event time and indicator should be independent from the observed censoring time ((*T,* Δ) **⊥** *C | A, W*) and given the observed covariates, the counterfactual event time and indicator should be independent from the observed treatment group for all intervention levels 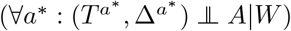. In particular, this implies that there is no unobserved confounding.
- *Consistency* If *A* = *a^*^*, then the observed and counterfactual event time and indicator should be aligned: 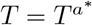 and 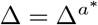 .

In Rytgaard & van der Laan’s [12] methodology, the counterfactual risks from the causal estimands (equations 3 and 4) are operationalized as estimates of the conditional cause-*j* Cumulative Incidence Function (CIF) *F_j_*(*t|a, w*) = ℙ(*T_j_ ≤ t|A* = *a, W* = *w*):

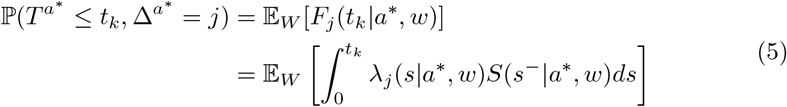

The CIF is computed by integrating point-wise risks over time up to a target time *t_k_*, where each point-wise risk is the product of a conditional cause-*j* hazard function

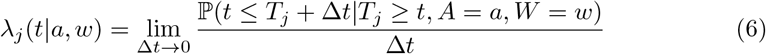

and a conditional event-free survival function

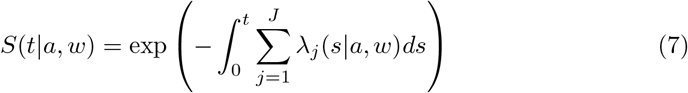

The conditional hazards denote the probability of experiencing event *j* in an infinitessemally small interval after a given time point, and the conditional event-free survival is the probability of having survived without encountering any event up to that time point.

### 2.2 Initial Estimation of Nuisance Parameters

Next to the cause-specific hazards *λ_j_*(*t|a, w*) for every target event *j* and the related event-free survival probability *S*(*t|a, w*), censoring survival *G*(*t|a, w*) and propensity scores *π*(*a|w*) need to be initially estimated in the first TMLE stage.

#### 2.2.1 Conditional Cause-specific Hazards, Event-free Survival and Censoring Survival

The initial estimation of the time-to-event nuisance functions is highly flexible in `PyTMLE`, and allows any model to be plugged in as long as it is implemented in a pycoxstyle class with methods `fit, predict surv`, and predict cif (competing risks) or predict cumulative hazards (no competing risks), and all inputs are in memory. Next to the two-dimensional standard input (which contains event time, event indicator, treatment group, and potentially tabular covariates), it is possible to pass additional inputs that can be higher-dimensional. Additionally, the package provides a wrapper for compatibility with models from `scikit-survival`.

For valid asymptotic inference with TMLE, it is important that the first-stage models have restricted complexity and do not overfit. Formally, all nuisance models should belong to a Donsker class [31]. If this cannot be guaranteed, the complexity condition can be weakened by estimation in a cross fitting [11] or super learning design [32], which we built into `PyTMLE` to support arbitrary ML models.

Super learners are designed to reduce variance by weighting the predictions of multiple base models into a final prediction [32]. The weights are chosen according to the base models’ loss in cross validation, where the loss function is utilized to evaluate the accuracy of the predictions. In the case of discrete super learners, only the predictions of the most accurate base learner are used, with all others receiving a weight of zero.

`PyTMLE` features a version of the state learner recently proposed by Munch & Gerds [20], a discrete super learner that can be used with any library of candidate survival or competing risks models. Unlike the discrete super learner implemented in `concrete`, it does not utilize a partial log-likelihood loss [33] and is thus not limited to Cox proportional hazards models. Additionally, it jointly evaluates combinations of models for event time and censoring distributions.

With observed inputs 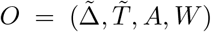 and based on the multi-state process 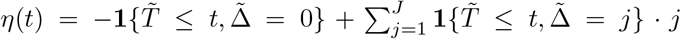, the loss function for a combination of event models and a censoring model is defined as the integrated Brier score 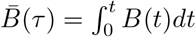, where *B*(*t*) is the Brier score [34] at time *t*:

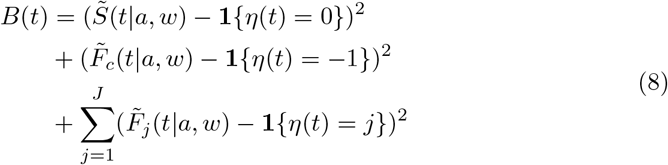

Here, 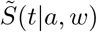 is the event-*and* censoring-free survival function:

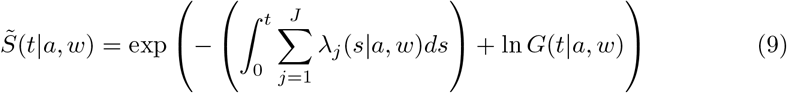

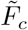 and 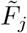 are the CIF for censoring or event *j*, respectively, defined as in equation 5 where *S* is replaced by 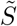.

By default, the `PyTMLE` super learner library contains a cause-specific Cox regression [35], a cause-specific random survival forest [2] (both from `scikit-survival`), and a vanilla instance of DeepHit [7] based on `pycox`. The state learner picks the combination of event and censoring models that has the smallest 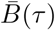 averaged over observations across all cross-validation folds. Compared to the state learner paper [20], our implementation is a simplification in that it always tests the same model for all *J* competing events, which decreases flexibility but can reduce the runtime for *J >* 1. Additionally, our super learner does not refit the best model combination on the whole data, so that there is still cross fitting if only a single base model candidate is given.

As an alternative to the default super learner or a super learner of a custom library of base models, it is also possible to pass pre-computed counterfactual initital estimates directly to the TMLE update step. This makes `PyTMLE` even more flexible and is useful especially for very advanced pipelines, e.g., involving hyperparameter tuning with nested cross validation or data loading from disk. Of course, the estimates should still come from models that are either Donsker or cross-fitted, ideally with super learning.

#### 2.2.2 Propensity Scores

If not pre-computed outside of `PyTMLE`, propensity scores are estimated by cross fitting the `StackingClassifier` from `scikit-learn` [36]. The model stacks the outputs of multiple base classifiers - including random survival forest [34] and gradient boosting classifiers [37] - and fits a meta-classifier to learn the weights of the individual base models for the final prediction. The `StackingClassifier` can be considered as a form of continuous super learning.

### 2.3 Targeted Update Step

The second stage of TMLE - the targeted update loop - operates on the initial estimates of the nuisance parameters from the first stage, pooled across all cross fitting folds. The update targets a set of EIC equations, with each equation corresponding to a specific combination of target event, target time, and intervention. Specifically, when dealing with *J* competing events, *K* time points and two interventions for the case of two treatment arms, the EIC becomes a *J × K ×* 2 vector *ϕ*. Each element in this vector pertains to the cause-specific risk of event *j* at time *t_k_* under the intervention *a^*^*. For tuple input 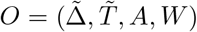 and the cause-specific counting process 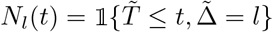, the elements of the EIC vector can be defined as

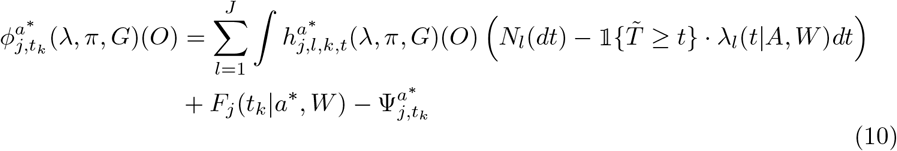

The EIC functions contain the “clever covariate”

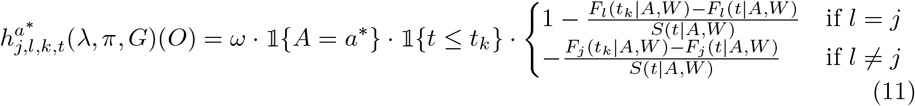

with nuisance weights

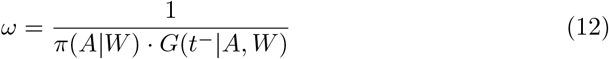

Exactly like in `concrete`, the cause-specific hazards *λ_j_* are updated in a loop with small recursive steps along a sequence of locally least favorable submodels:

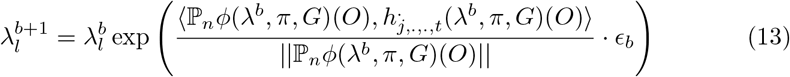

The step size *ϵ_b_* is halved whenever at step *b*

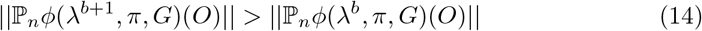

until the convergence criterion is fulfilled at iteration *B*:

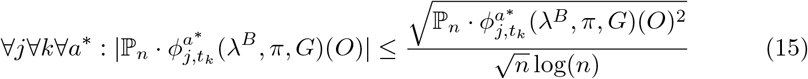

### 2.4 Confidence Intervals

Like `concrete`, `PyTMLE` offers CIs based on the closed-form standard errors derived from the EIC. Under asymptotic normality, the CIs of effect estimates are computed as 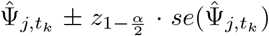. Unfortunately, it has been observed that these CIs can have a poor coverage if the positivity assumption is violated or events are rare [38]. Therefore, our package has an alternative option to compute CIs with a non-parametric bootstrap (potentially stratified by event). As recommended in the literature [38, 39], we bootstrap only the second TMLE stage and keep the initial estimators fixed across all bootstrap iterations. This not only makes each iteration faster, but also allows bootstrapping to be applied even if some or all nuisance parameters are precomputed outside of the package. The final bootstrap distribution is generated only from iterations that reached the convergence criterion (equation 15). CIs can be set to the 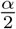 and 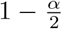 quantiles of the bootstrapped distribution of counterfactual risk or ATE estimates.

### 2.5 Sensitivity Analysis and Checks of Assumptions

Even though violations of the conditional exchangeability assumption can never be ruled out completely in real-world data, there are ways to assess how robust RD and RR estimates are with regard to unmeasured confounding. One of them is the *E-value*, which corresponds to the minimum association level that any unobserved confounder would need to have to completely nullify a predicted treatment effect on the outcome, conditional on all measured covariates [30].

It is very straightforward to compute E-values for the ATE defined in terms of RR (equation 4):

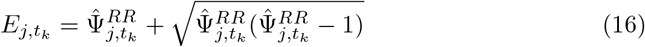

if 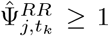, or substituting 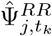 with 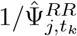 otherwise. E-values are typically not only reported for the point estimate, but also for the CI limit closer to the null (1 for RRs).

As Van der Weele & Ding [30] showed, RR-based point estimates can be approximated from an RD-based point estimate, in our case with the simple transformation 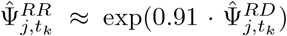. For CI E-values, bounds can either be re-computed via 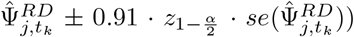 from EIC-derived standard errors, or by transforming bootstrapped quantiles exactly like the point estimates.

Since E-values are on an arbitrary RR scale and cannot be interpreted without contextualization, we integrated a benchmark for observed covariate E-values as proposed by McGowan & Greevy [40]. Here, the complete TMLE procedure (including both the first and second stage) is refitted on data from which one observed covariate is dropped at a time. Observed covariate E-values are then computed based on the CIs of the full and the reduced models to get a reference for the interpretation of the E-values predicted for unmeasured confounders.

Our implementation of E-values and the benchmark was adapted from the dowhy package [41, 42].

Like concrete, our package also has an option to plot nuisance weights *ω* (equation 12) for specified target times and target events to detect positivity violations.

## 3 Demonstration

To demonstrate the functionality of PyTMLE, we applied the package to the Hodgkin’s lymphoma dataset which is available in the R package `randomForestSRC` [43] and described in Pintilie [44]. The dataset includes longitudinal follow-up of patients diagnosed with early-stage Hodgkin’s lymphoma, who received either radiation therapy alone or additional chemotherapy.

Patients were followed for the occurrence of two competing events: Event 1 (relapse) and Event 2 (death). Censoring occurred for patients lost to follow-up or event-free at the end of the study period. The dataset includes several baseline covariates commonly used for risk adjustment, including age at diagnosis, sex, disease stage, and histological subtype.

Using `PyTMLE`, we estimated the causal effect of treatment on the cumulative risk of each event type at specified target times, alongside diagnostic outputs such as nuisance weight distributions and E-values to assess positivity and robustness.

The following code demonstrates how to initialize a `PyTMLE` estimator and fit it to the Hodgkin’s disease dataset, assuming that the “status” column is coded as `0=censored, 1=relapsed, 2=deceased` and including repeated model re-fitting for E-value benchmarking:

**Listing 1** Initializing and fitting the PyTMLE model

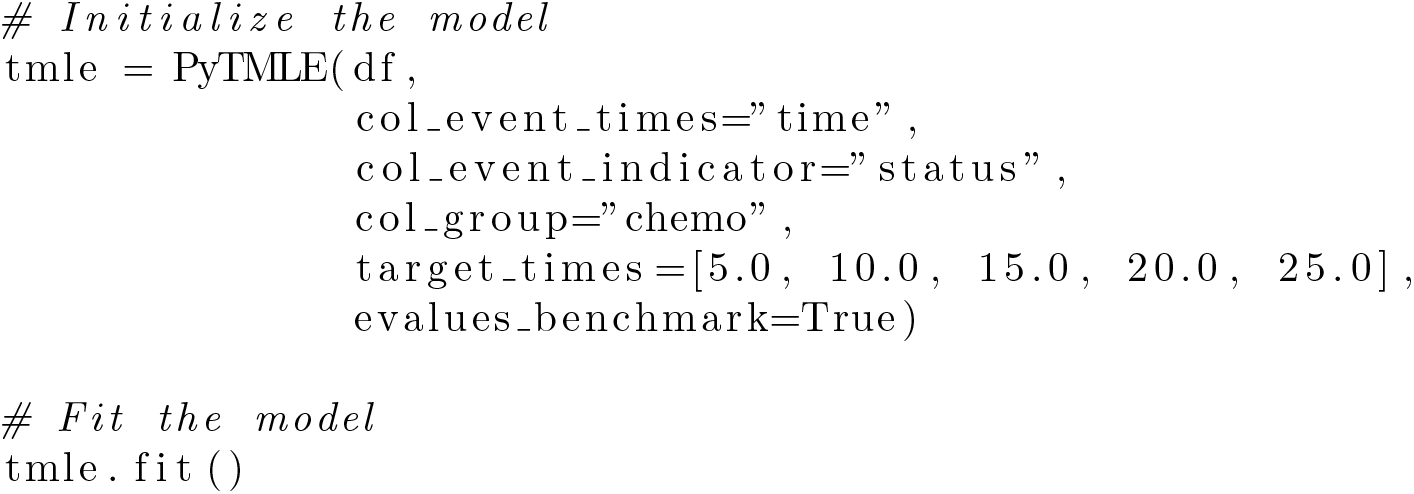

### 3.1 Risk Estimates Over Time

The top half of Figure 1 (generated with tmle.plot()) shows the predicted counterfactual CIF for both Event 1 and Event 2 over time. TMLE estimates are plotted with 95% CIs based on the EIC, and G-computation estimates are included for reference.

**Fig. 1.**
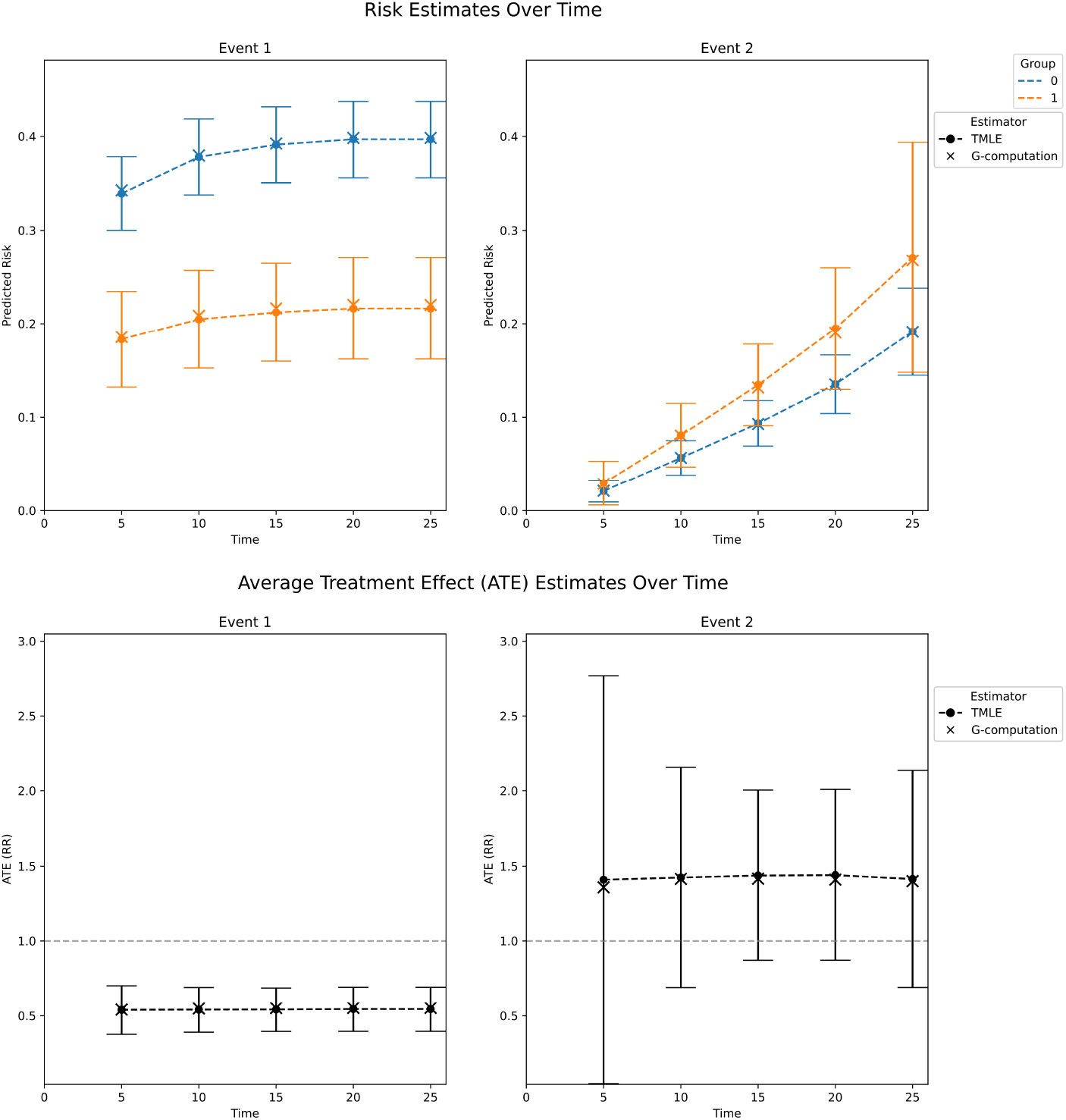
TMLE predictions. Top: Predicted risk estimates over time for Event 1 (relapse) and Event 2 (death) using TMLE and G-computation. TMLE includes 95% CIs. For both events, TMLE and G-computation yield similar trends, though treatment effects differ by event type. Bottom: ATE estimates (risk ratio scale) over time for Events 1 and 2. TMLE indicates a persistent protective effect for Event 1 (*RR <* 1), and no effect for Event 2 (CIs cover 1.0). G-computation estimates are close those updated with TMLE.

- For Event 1 (relapse), Group 0 (blue) consistently shows a higher risk than Group 1 (orange), indicating a potential protective effect of the treatment. Both estimators yield similar risk trajectories over time, with TMLE typically providing slightly higher estimates.
- For Event 2 (death), risk increases more gradually and diverges over time. The gap between groups widens over time (particularly from time 15 onward), but CIs keep overlapping across all targeted times.

### 3.2 Average Treatment Effect (ATE) Estimates

The bottom half of Figure 1 (generated with tmle.plot(type=“rr”)) displays the ATE estimates on the RR scale over time for both Event 1 (relapse) and Event 2 (death).

- For Event 1, the CIs of TMLE ATE estimates remain consistently below 1.0, indicating a sustained protective effect of chemotherapy on relapse risk over time. There is not a large difference from the effects estimated via G-computation in the first place.
- In contrast, for Event 2, both TMLE and G-computation estimates are above 1.0 at all time points, but TMLE CIs consistently cover 1.0. This indicates no effect of chemotherapy on mortality.

These results suggest that while chemotherapy may reduce the risk of relapse, it does not seem to impact the risk of death.

### 3.3 Positivity Diagnostics

To assess if the positivity assumption is met, we examined the distribution of nuisance weights defined as *π*(*a | w*) *· G*(*t | a, w*) at two key time points.

- At time 5.0 (left half of Figure 2, `tmle.plot nuisance weights(time=5.0)`), the distributions are bounded away from zero for both treatment groups. This suggests that the positivity assumption is reasonably well met early in the time horizon.
- At time 25.0 (right half of Figure 2, `tmle.plot nuisance weights(time=25.0)`), both groups exhibit a strong concentration of weights near zero, particularly Group 0. This indicates potential lack of support for this target time due to a drastic reduction of censoring survival probabilities for all observations in the analyis, which could lead to unstable estimates and inflated variances in the TMLE procedure.

**Fig. 2.**
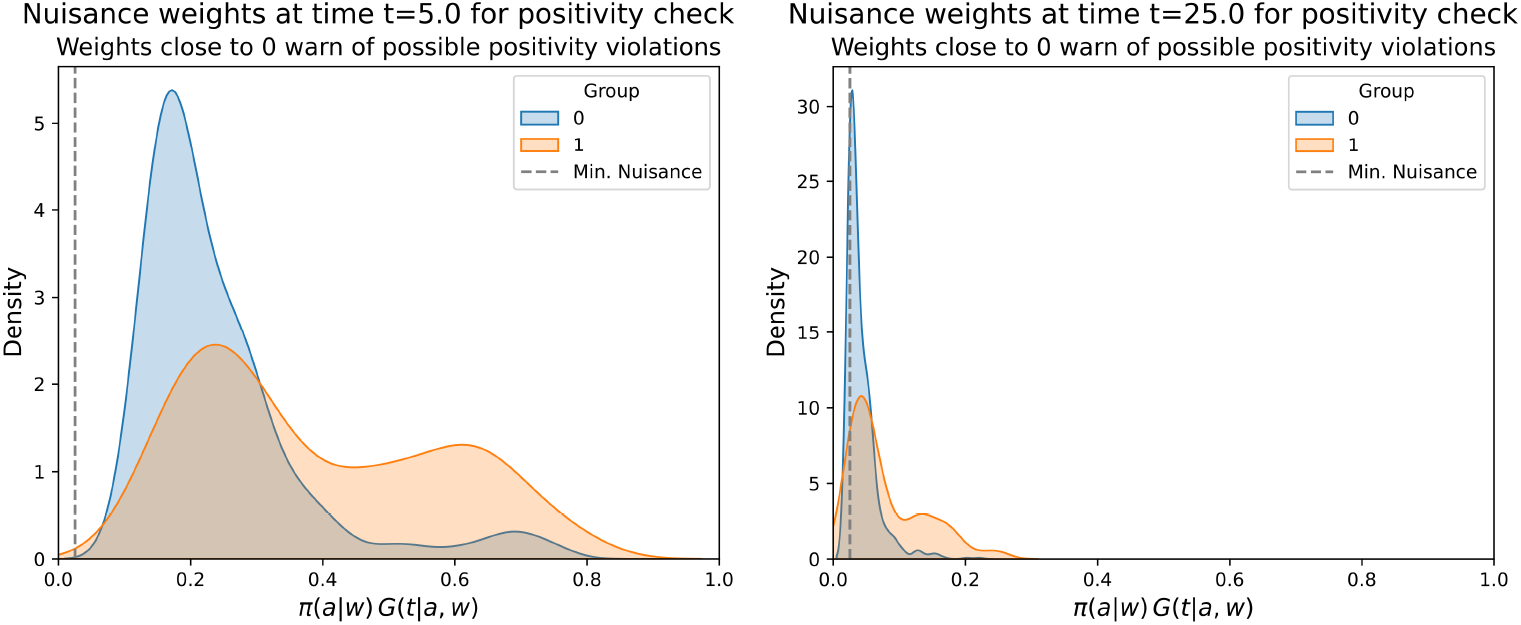
Distribution of nuisance weights at time 5.0 (left) and 25.0.

Monitoring these weights over time is essential to detect regions of poor support and to motivate a modification of target times if needed.

### 3.4 Sensitivity Analysis: E-values

Figure 3 presents an E-value contour plot for Event 1 at time 25.0 (`tmle.plot evalue contours(type=“rr”, time=25.0, event=1)`), quantifying how robust the observed treatment effect is to potential unmeasured confounding.

**Fig. 3.**
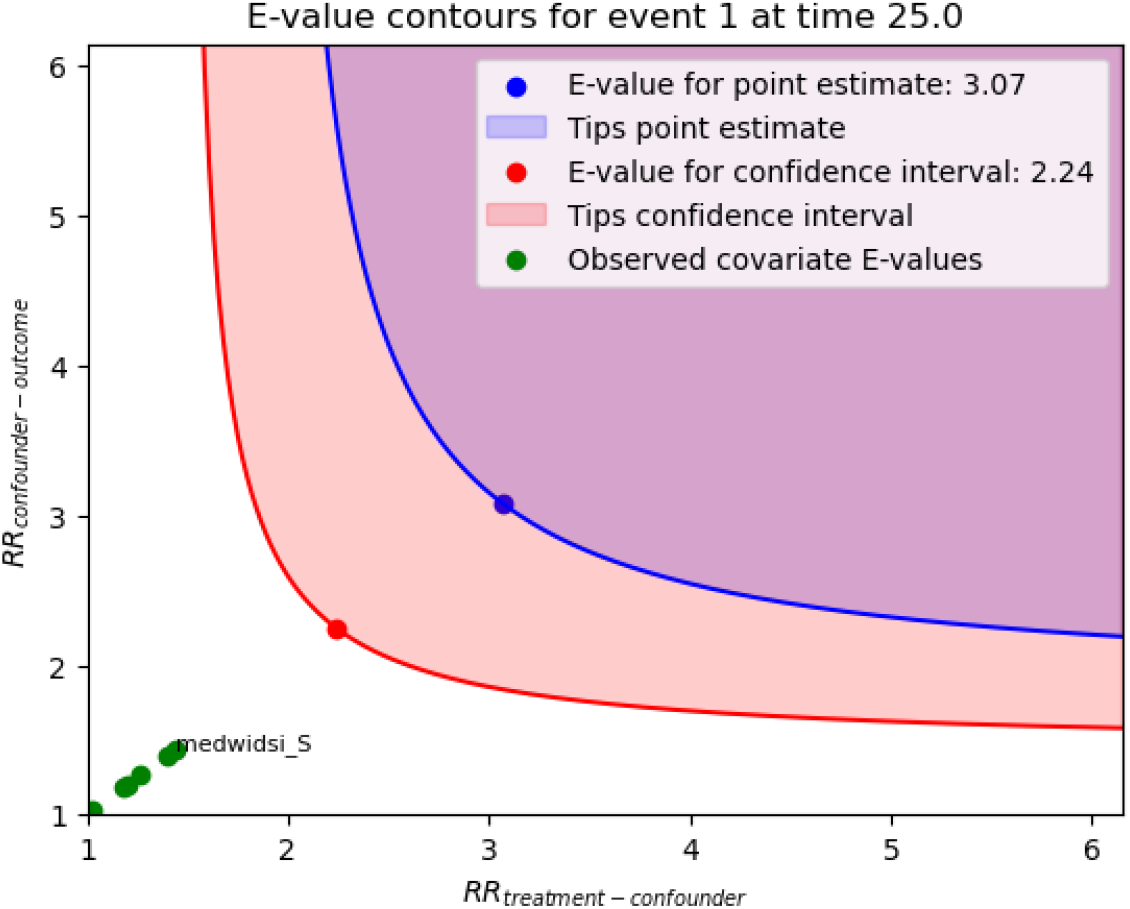
E-value contour plot for Event 1 at time 25.0. Blue and red curves indicate the E-values for the point estimate (3.07) and confidence interval lower bound (2.24), respectively. Observed covariates (green points) show weaker associations (strongest for variable “small mediastinum involvement”), indicating that the effect estimate is robust to known sources of confounding.

- The E-value for the point estimate is 3.07, meaning that an unmeasured confounder would need to have a risk ratio association of at least 3.07 with both the treatment and the outcome to fully explain away the observed effect.
- The E-value for the bound of the confidence interval closer to the null is 2.24, which means that the bound would only tipped by a confounder of at least this association strength with treatment and outcome.
- Green dots represent benchmark E-values computed for observed covariates, all of which fall below the red and blue contours. This suggests that an unobserved confounder would need an association strength with treatment and outcome that is clearly above that connected to the observed covariates, which makes it unlikely that such a confounder exists.

These results underscore the robustness of the effect estimate for Event 1 at time 25.0 against unmeasured confounders of moderate strength.

## 4 Conclusions

We developed the Python package `PyTMLE` for targeted estimation of treatment effects on time-to-event and competing risks outcomes from observational data. By providing a fully flexible first TMLE stage for the estimation of nuisance parameters, the package exploits the pairing of desirable statistical properties with data-adaptive estimation to a larger extent than previously released implementations did. Sufficiently fast convergence of the first stage is supported by super learning, which is based on the *state learner* for the survival-related functions. Technically, `PyTMLE` currently supports models from `scikit-survival` and neural network models defined via the pycox framework. In practical applications, the robustness of predictions and positivity can be assessed with two alternative techniques for CIs, E-value benchmarks, and plots of nuisance weights. We demonstrated the usability of the package on an established clinical dataset from the competing risks literature.

While pycox offers the most comprehensive set of deep learning survival models currently available, it covers only a fraction of those cataloged by Wiegrebe et al. [45]. Moreover, its inability to load data from external storage restricts its utility for complex datasets like images. Future Python packages that better integrate deep learning with survival analysis may be considered for inclusion in `PyTMLE`.

Currently, the package only supports baseline confounders and point treatments. Potential future developments could be extensions to time-varying confounders and treatment plans.

We hope that PyTMLE encourages ML practitioners - especially in fields like biomedical data science and epidemiology - to apply the powerful framework of targeted estimation for treatment effects on survival outcomes, even on data that is high-dimensional and potentially multimodal.

## Availability and Requirements

- **Project name**: PyTMLE
- **Project home page**: https://github.com/SCAI-BIO/pytmle
- **Operation system(s)**: Platform independent
- **Programming language**: Python
- **Other requirements**: Python 3.9 or higher
- **License**: Apache-2.0
- **Any restrictions to use by non-academics**: No

### List of Abbreviations

*AI*: Artificial Intelligence
*ATE*: Average Treatment Effect
*CI*: Confidence Interval
*CIF*: Cumulative Incidence Function
*EIC*: Efficient Influence Curve
*ML*: Machine Learning
*RD*: Risk Difference
*RR*: Risk Ratio
*TMLE*: Targeted Maximum Likelihood Estimation

## Declarations

### Ethics approval and consent to participate

Not applicable.

### Consent for publication

Not applicable.

### Data availability

The Hodgkin’s lymphoma dataset analyzed during the current study is available alongside the code in the PyTMLE repository, https://github.com/SCAI-BIO/pytmle/tree/main/docs/source/notebooks.

### Author contribution

Conceptualization, methodology, supervision, project administration, and funding acquisition: HF. Data curation, formal analysis, visualization, investigation, validation, and writing—original draft: JG, MA. Writing—review and editing: JG, MA and HF. All authors contributed to the article and approved the submitted version.

## Acknowledgements

On behalf of COMMUTE and Real4Reg, we would like to express our gratitude for the support and collaboration received throughout these projects.

